# Progression of daily-life tremor measures in early Parkinson disease: a longitudinal continuous monitoring study

**DOI:** 10.64898/2025.12.23.25342892

**Authors:** Nienke A. Timmermans, Ioan Gabriel Bucur, Diogo C. Soriano, Erik Post, Hayriye Cagnan, Sooyoon Shin, Max A. Little, Yordan P. Raykov, Bastiaan R. Bloem, Rick C. Helmich, Luc J.W. Evers

**Author notes:** Corresponding author: Nienke A. Timmermans.

## Abstract

**Background:** Sensitive outcome measures are critical for evaluating the efficacy of novel treatments for Parkinson disease (PD). Recently, we demonstrated that a tremor detection algorithm could reliably detect and quantify real-life PD tremor from wearable sensor data. Here, we assess the sensitivity to change of sensor-derived daily-life tremor measures over two years in an unmedicated and medicated cohort of persons with early PD.

**Methods:** We used two-year continuous wrist sensor data (median wear time: 22 hours/day) from the Personalized Parkinson Project (n=462 medicated; n=78 unmedicated at baseline), in combination with annual clinical evaluations of tremor severity. From the raw gyroscope data, we derived previously validated weekly measures for tremor time and power, which were smoothed over time using piecewise linear trend estimation. One- and two-year standardized response means (SRMs) were computed to compare the sensitivity to change between the sensor-derived tremor measures and clinical tremor scores.

**Findings:** In unmedicated participants with tremor, sensor-derived tremor measures demonstrated a high sensitivity to progression (two-year SRMs ranged from 0.67 to 1.09), which was significantly larger than clinical tremor scores (two-year SRMs ranged from 0.21 to 0.41). In medicated participants, sensor-derived tremor time decreased (two-year SRM of -0.18 in participants with tremor), which was associated with both an increase in dopaminergic medication dose and higher disease duration. In contrast, the sensor-derived tremor power measures and clinical rest tremor scores (measured in the OFF state) increased slightly (two-year SRMs ranging from 0.11 to 0.27).

**Interpretation:** Prior to initiation of symptomatic treatment, sensor-derived daily-life tremor measures are substantially more sensitive to disease progression than clinical tremor scores, making them a promising tool to evaluate the efficacy of disease-modifying treatments in early PD.

## Introduction

As the prevalence of Parkinson disease (PD) continues to increase^1^, disease-modifying or more effective symptomatic treatments are highly warranted to alleviate the burden of this chronic and progressive disease. However, adequate measures of clinical disease progression are still lacking, making it difficult to evaluate the effectiveness of novel treatments^2,3^. The currently used Movement Disorder Society - Unified Parkinson’s Disease Rating Scale (MDS-UPDRS) depends on episodic and subjective assessments, limiting its sensitivity to measure disease progression^4^. Wearable sensors that continuously and objectively assess PD symptoms in natural environments could address these issues^5–8^.

In addition to being sensitive to change, digital outcome measures should capture concepts that are meaningful to patients^9,10^. In early PD, tremor is experienced as one of the most bothersome symptoms^11–13^. Tremor not only interferes with daily activities, but has sensory and psychosocial impacts as well^14^. It is influenced by emotional and cognitive stress, voluntary movements, and timing of treatments, making it a highly fluctuating symptom^15^. This complicates its evaluation during the typically brief, episodic in-person clinical visits^11^.

Earlier studies have shown that PD tremor can be measured accurately with wrist-worn sensors^16^. However, longitudinal studies evaluating tremor progression in daily-life are scarce, with only one study with a one-year follow-up^7^. Therefore, the ability of sensor-derived tremor measures to capture tremor progression remains largely unknown.

Furthermore, previous studies investigating tremor progression using MDS-UPDRS assessments have yielded conflicting results in varying study settings: PD tremor could increase, remain stable, or decrease over time^17–20^. Investigating tremor progression in naturalistic environments could improve our understanding of tremor pathophysiology.

In this study, we examined tremor progression over two years using continuous free-living sensor data from 540 early-stage PD participants from the Personalized Parkinson Project^21,22^. We have previously developed and validated an open-source algorithm that enables reliable monitoring of PD tremor in real-life conditions^23^. In this study, we compare the sensitivity to progression of the sensor-derived measures against clinical tremor observations and patient-reported tremor severity based on the MDS-UPDRS. To isolate the effects of disease progression and symptomatic medication, we separately analyze participants who remained unmedicated throughout the study follow-up and those receiving symptomatic drug therapy. Additionally, we assess the effect of changes in dopaminergic medication dose on sensor-derived tremor measures. Collectively, this study aims to lay the foundation for using real-life, objective tremor outcomes to measure disease progression, both in clinical trials evaluating disease-modifying therapies and in cohort studies aiming to increase our understanding of tremor pathophysiology.

## Methods

### Study design and participants

Data were obtained from the Personalized Parkinson Project (PPP) cohort and the PPP De Novo cohort^21,22^. The PPP cohort included 520 subjects with early PD (disease duration ≤ 5 years). In the PPP De Novo cohort, 103 recently diagnosed (disease duration ≤ 2 years) and treatment-naïve PD patients were included, who were not expected to start treatment in the first year after inclusion. In both cohorts, participants were monitored continuously for two years using the Verily Study Watch (median wear time of 22 hours/day^24^), collecting raw gyroscope, accelerometer and photoplethysmography data. In addition, in-clinic MDS-UPDRS assessments were performed at baseline and after one and two years (see Figure 1).

**Figure 1:**
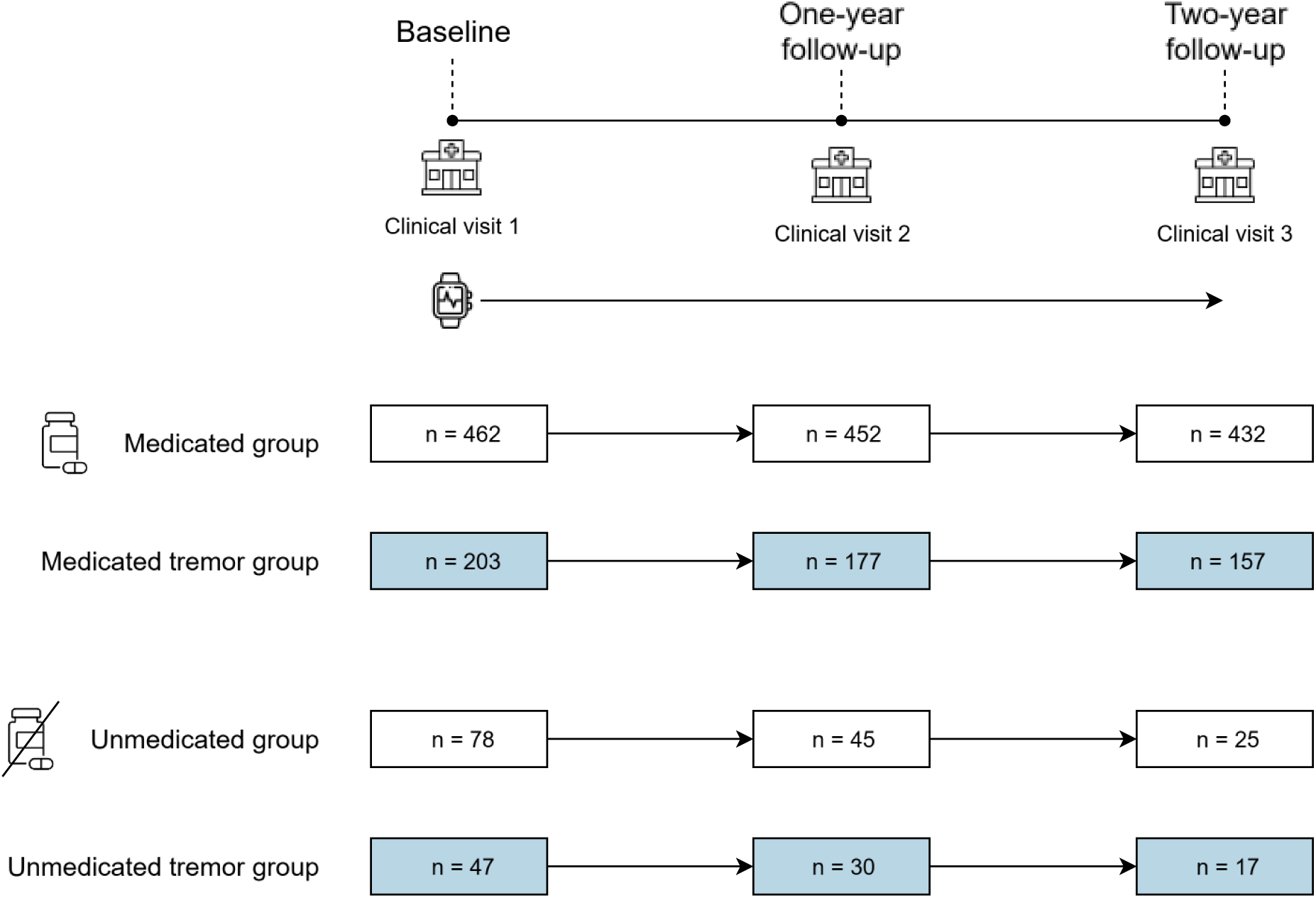
Schematic overview of the study design of the Personalized Parkinson Project and the subgroups used in this study. The tremor groups contain participants for whom the amount of detected tremor exceeded the false positive threshold at baseline and at follow-ups. The number of included participants decreased over time due to dropout, initiation of symptomatic drug treatment (in the unmedicated groups), or disappearance of tremor (in de tremor groups). The subgroups used in our primary analysis focusing on long-term tremor progression are highlighted in blue.

Participants wore the watch on their preferred side. We excluded participants that switched their watch side during the study period (n= 25), and participants with fewer than eight weeks of sensor data (n=22). Other exclusion criteria were having an alternative diagnosis (n=15), or having an unknown start date of symptomatic medication (n=21). This resulted in the inclusion of 540 participants with PD.

Because of the effects of symptomatic medication on tremor, for our primary analysis focusing on long-term tremor progression we divided participants into: 1) a medicated group with participants who had already initiated symptomatic drug treatment at baseline; and 2) an unmedicated group, consisting of participants who were untreated at the time of inclusion. The number of participants in the unmedicated group decreased over time due to initiation of treatment during the follow-up period (Supplementary Figure 1). Within these two groups, we created additional subgroups only including participants with tremor. This was based on whether the amount of watch-detected tremor exceeded the false positive threshold of 3.5% tremor time, which was the 90^th^ percentile of tremor time in controls^23^.

Together, this resulted in four analysis groups: medicated, medicated tremor, unmedicated, and unmedicated tremor, with group sizes varying across follow-up (Figure 1).

The PPP studies were conducted in accordance with the Declaration of Helsinki and Good Clinical Practice guidelines and was approved by the local medical ethics committee (Commissie Mensgebonden Onderzoek, regio Arnhem-Nijmegen, reference number 2016–2934 for PPP and 2020-6111 for PPP de Novo). All participants provided informed consent prior to enrollment.

### Sensor-derived tremor measures

The continuous, high-frequency (i.e., 100 Hz) gyroscope data collected by the watch was processed using the ParaDigMa toolbox^25^, which contains a previously validated algorithm to detect and quantify PD rest and re-emergent tremor^23^. Three weekly aggregated tremor measures were derived: tremor time, modal tremor power, and the 90^th^ percentile of tremor power. Tremor time was calculated as the percentage of inactive daytime with detected tremor. The modal tremor power captures the typical tremor severity, whereas the maximal tremor severity is represented by the 90^th^ percentile of tremor power. To reduce computational load and data volume, we derived the weekly tremor measures every other week. Furthermore, the tremor power measures were only derived for weeks in which the amount of detected tremor time exceeded the false positive threshold, to avoid tremor power estimates in weeks where the majority of detected tremor likely reflected false positives. Further details of the tremor detection algorithm are described elsewhere ^23^.

To focus on long-term tremor progression rather than short-term fluctuations or measurement error, we used piecewise linear trend estimation (L1-trend filtering) to smooth the biweekly tremor measures over the two-year measurement period^26,27^. The trend was obtained by minimizing the following function:

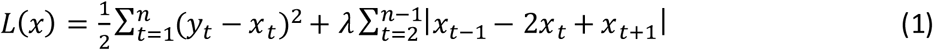

where 𝑦_𝑡_is the weekly tremor measure and 𝑥_𝑡_ is the fitted trend at time *t*. For each tremor measure, we determined the optimal smoothing parameter 𝜆 by selecting the value which yielded the best predictive accuracy of randomly masked observations across participants (i.e., minimizing the mean absolute residual error across all participants, using leave-one-point-out cross-validation). The optimal 𝜆 and the proportion of variance explained by the fitted trends per measure are given in Supplementary Table 1. We inter- and extrapolated the fitted trend for all weeks for which there was a tremor measure available in the surrounding four weeks. The resulting trends were used for all subsequent analyses.

### Sensitivity to tremor progression

We define tremor progression as long-term changes in tremor time and power, irrespective of their underlying cause (e.g., neurodegeneration, symptomatic treatment or behavior). The sensitivity to tremor progression of sensor-derived measures was assessed in the medicated and unmedicated groups separately, and compared to the sensitivity of MDS-UPDRS rest tremor scores and patient-reported tremor score (items 3.17, 3.18 and 2.10), using one- and two-year standardized response means (SRMs). An SRM of 0.2-0.5 indicates small responsiveness, 0.5-0.8 moderate responsiveness, and >0.8 large responsiveness^28^.

95% confidence intervals were constructed using 10,000 bootstrap samples. Week 2 of the study period was selected as the baseline week to exclude potential effects of the baseline study visit. For the two-year follow-up measures, week 100 was chosen to maximize the sample size available. Similarly, week 50 was used as the one-year follow-up week. Clinical data were only included if the visit date was at 50±10 weeks from baseline for the second visit, and at 100±20 weeks from baseline for the third visit. Before assessing the SRM of tremor time, we transformed this measure using the logit function to approximate normality.

### Adjustment for censoring

To assess disease progression without the effects of symptomatic medication, we selected participants that remained untreated during the one- or two-year follow-up period. However, this could introduce selection bias due to informative censoring, for example if participants with more tremor initiate treatment earlier. To account for this, we applied inverse probability-of-censoring weighting (IPCW), which reweighs remaining uncensored participants to reflect the composition of the cohort at baseline^29^. We estimated the effect of tremor on the probability of censoring due to treatment initiation by fitting a Cox proportional hazards model in participants that were unmedicated at baseline (n=78).

Tremor time was used as the time-varying covariate^30^, averaged over two measurement weeks. Different lags (4, 6 and 8 weeks) between the measurement of tremor and the initiation of treatment were evaluated in separate models, since we expected that there is a delay between worsening of tremor and the moment of treatment initiation. We subsequently used the model with the best overall fit (highest concordance statistic) to estimate the probability of each participant to remain unmedicated at each follow-up week. The inverse probabilities were then used to reweigh participants within the unmedicated group for each week.

### Sensitivity analyses

To assess the robustness of our findings, we performed several sensitivity analyses. We assessed the effect of follow-up duration and smoothing on the observed SRMs, by visualizing SRMs over time based on smoothed and unsmoothed data. Furthermore, we studied the effect of IPCW on the results by comparing the weighted and unweighted SRMs in the unmedicated group. Lastly, to assess the difference in tremor progression between the medicated and unmedicated groups while excluding potential effects of age, disease duration and watch position (whether the watch was worn on the more- or less-affected side^31^), we matched these subgroups at baseline using propensity score matching. We estimated propensity scores using logistic regression and matched participants 1:1 using nearest-neighbor matching without replacement. The more- or less-affected side was based on the sum of MDS-UPDRS III tremor scores on both sides (4 items) in the OFF state. If those scores were equal, the patient-reported more-affected side was selected. If the participant also reported equally affected sides, we chose the side with the larger total MDS-UPDRS III OFF score.

### Correlation with changes in patient-reported tremor score

To assess how objectively measured tremor changes relate to experienced tremor burden, we correlated the two-year changes in sensor-derived tremor measures to the changes in the patient-reported tremor score (MDS-UPDRS 2.10). For comparison, we also assessed how changes in the clinical evaluation of rest tremor ( MDS-UPDRS 3.17 and 3.18) correlate to the patient-reported tremor score.

### Effect of changes in dopaminergic medication dose

To study the effect of changes in dopaminergic medication dose on daily-life tremor measures, we performed two additional analyses. First, we assessed the relation between two-year sensor-derived tremor progression and changes in levodopa equivalent daily dose (LEDD), corrected for disease duration at baseline and watch position (more- or less-affected side), by using multivariable linear regression in the medicated group. Secondly, we assessed the responsiveness of the sensor-derived tremor measures to the initiation of dopaminergic treatment, using participants that initiated dopaminergic medication during the study period (n=49). We calculated the absolute changes between the week before initiation of treatment and each week in a period of 0 to 14 weeks since the initiation of treatment, and we derived SRMs from these changes.

## Results

Table 1 shows the demographic and clinical characteristics, as well as sensor-derived tremor measures of the medicated and unmedicated group at baseline.

**Table 1:**
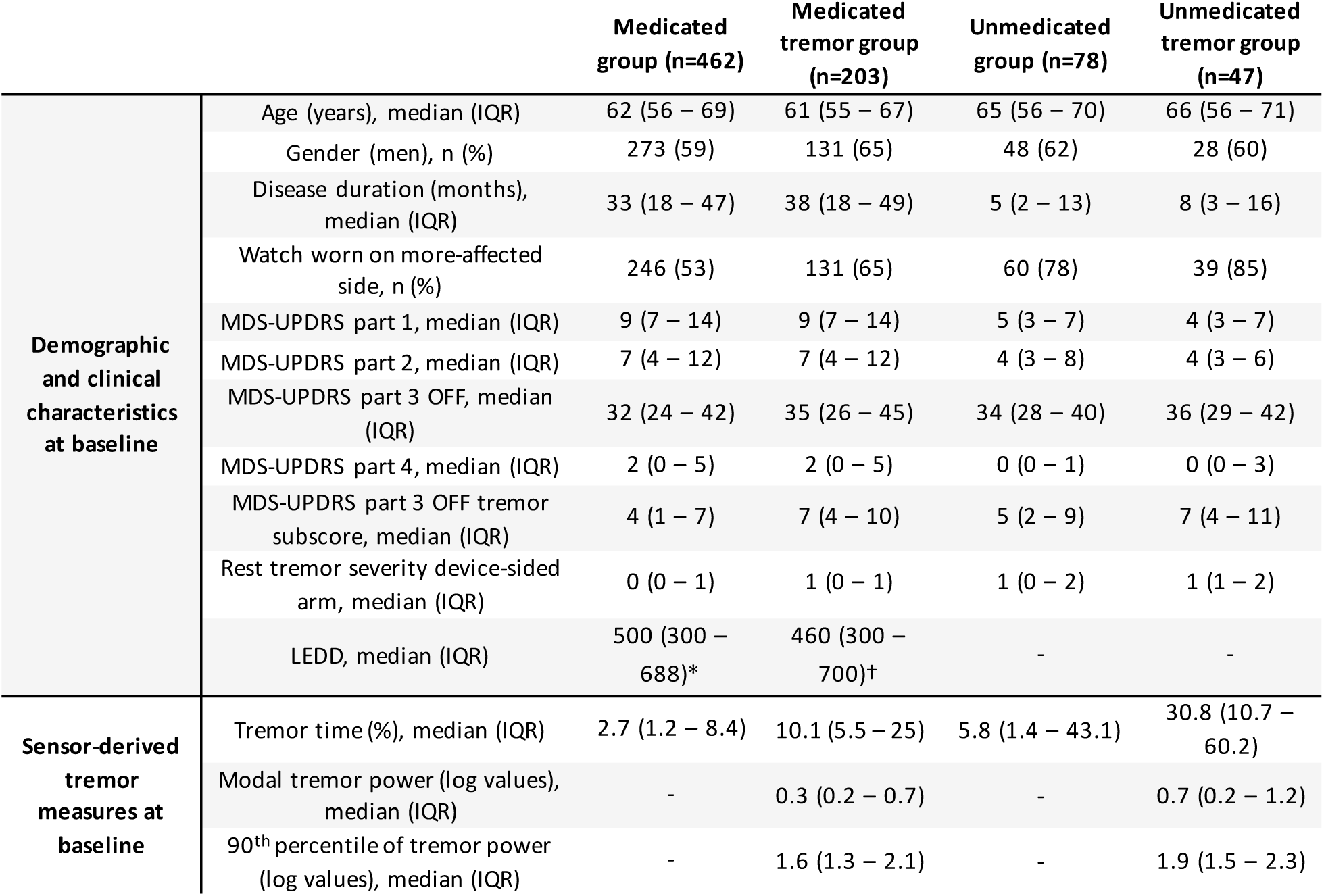
Demographic and clinical characteristics and sensor-derived tremor measures at baseline of the medicated and unmedicated groups at baseline. The modal and 90^th^ percentile of tremor power were assessed in all participants with tremor time above the false positive threshold at baseline (n=203 for the medicated group and n=47 for the unmedicated group). IQR: inter-quartile range. LEDD: Levodopa Equivalent Daily Dose. MDS-UPDRS: Movement Disorder Society-Sponsored Revision of the Unified Parkinson’s Disease Rating Scale. Part 1: non-motor experiences of daily living. Part 2: motor experiences of daily living. Part 3: motor examination. Part 4: motor complications. * LEDD information of 20 participants was incomplete and 13 participants were using non-dopaminergic medication. † LEDD information of 12 participants was incomplete and 9 participants were using non-dopaminergic medication.

|  |  | Medicated group (n=462) | Medicated tremor group (n=203) | Unmedicated group (n=78) | Unmedicated tremor group (n=47) |
| --- | --- | --- | --- | --- | --- |
| Demographic and clinical characteristics at baseline | Age (years), median (IQR) | 62 (56 – 69) | 61 (55 – 67) | 65 (56 – 70) | 66 (56 – 71) |
|  | Gender (men), n (%) | 273 (59) | 131 (65) | 48 (62) | 28 (60) |
|  | Disease duration (months), median (IQR) | 33 (18 – 47) | 38 (18 – 49) | 5 (2 – 13) | 8 (3 – 16) |
|  | Watch worn on more-affected side, n (%) | 246 (53) | 131 (65) | 60 (78) | 39 (85) |
|  | MDS-UPDRS part 1, median (IQR) | 9 (7 – 14) | 9 (7 – 14) | 5 (3 – 7) | 4 (3 – 7) |
|  | MDS-UPDRS part 2, median (IQR) | 7 (4 – 12) | 7 (4 – 12) | 4 (3 – 8) | 4 (3 – 6) |
|  | MDS-UPDRS part 3 OFF, median (IQR) | 32 (24 – 42) | 35 (26 – 45) | 34 (28 – 40) | 36 (29 – 42) |
|  | MDS-UPDRS part 4, median (IQR) | 2 (0 – 5) | 2 (0 – 5) | 0 (0 – 1) | 0 (0 – 3) |
|  | MDS-UPDRS part 3 OFF tremor subscore, median (IQR) | 4 (1 – 7) | 7 (4 – 10) | 5 (2 – 9) | 7 (4 – 11) |
|  | Rest tremor severity device-sided arm, median (IQR) | 0 (0 – 1) | 1 (0 – 1) | 1 (0 – 2) | 1 (1 – 2) |
|  | LEDD, median (IQR) | 500 (300 – 688)* | 460 (300 – 700)† | - | - |
| Sensor-derived tremor measures at baseline | Tremor time (%), median (IQR) | 2.7 (1.2 – 8.4) | 10.1 (5.5 – 25) | 5.8 (1.4 – 43.1) | 30.8 (10.7 – 60.2) |
|  | Modal tremor power (log values), median (IQR) | - | 0.3 (0.2 – 0.7) | - | 0.7 (0.2 – 1.2) |
|  | 90<sup>th</sup> percentile of tremor power (log values), median (IQR) | - | 1.6 (1.3 – 2.1) | - | 1.9 (1.5 – 2.3) |

### Sensitivity to tremor progression

Tremor progression was defined as long-term, within-subject changes in tremor time or power. Figure 2 shows these changes at the group level, while Figure 3 provides examples of typical individual progression patterns. The distribution of absolute sensor-derived tremor measures over time is given in Supplementary Figure 2.

**Figure 2:**
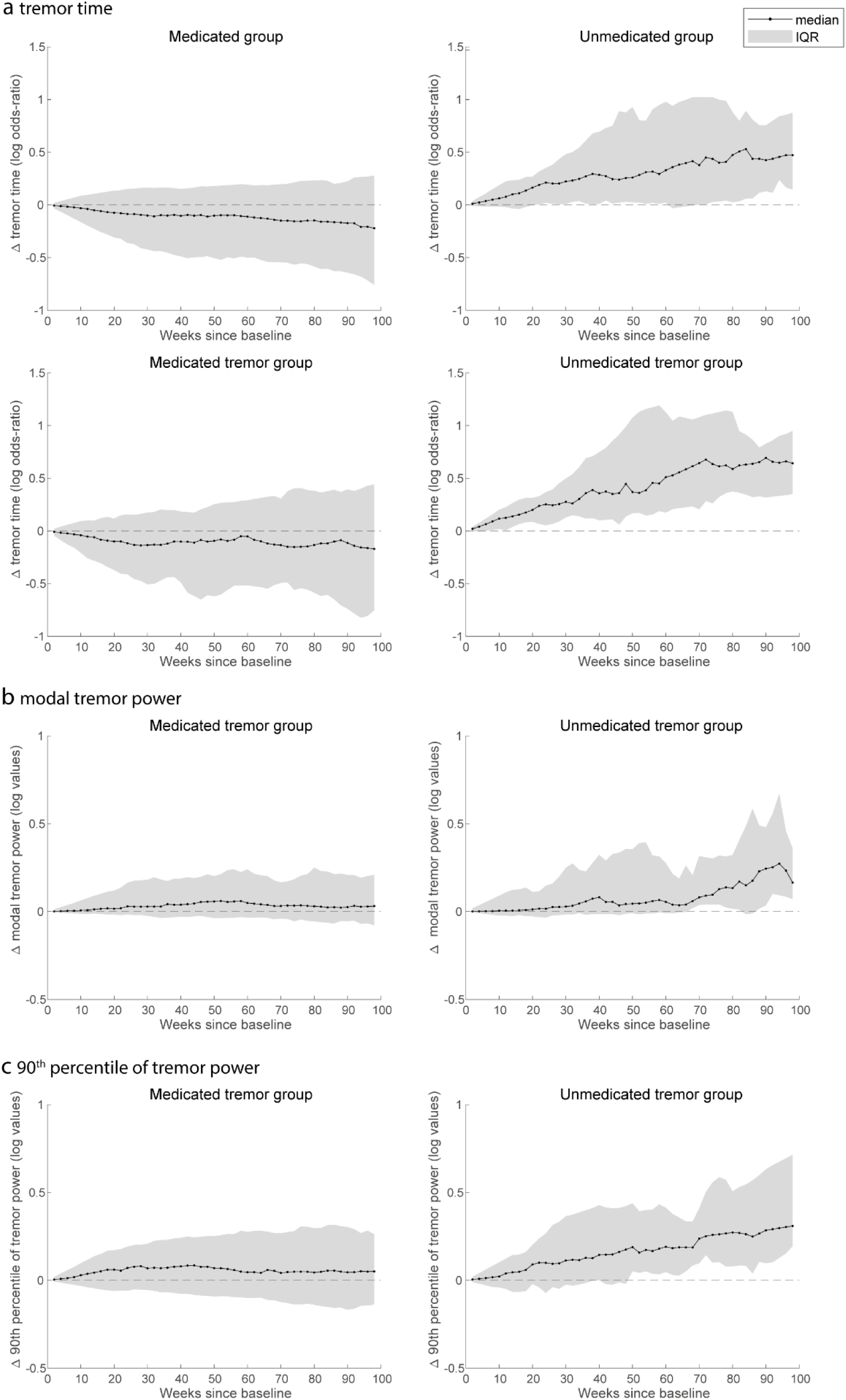
Median (and IQR of) absolute within-subject change since baseline of sensor-derived weekly tremor measures, observed in the medicated (on the left) and unmedicated group (on the right). Tremor time (a) was derived for all participants for whom data was available at baseline and at follow-up (upper plots), and subsequently for all participants with tremor time above the false positive threshold at baseline and at follow-up (lower plots). The modal (b) and 90^th^ percentile of tremor power (c) were also assessed for all participants with tremor time above the false positive threshold.

**Figure 3:**
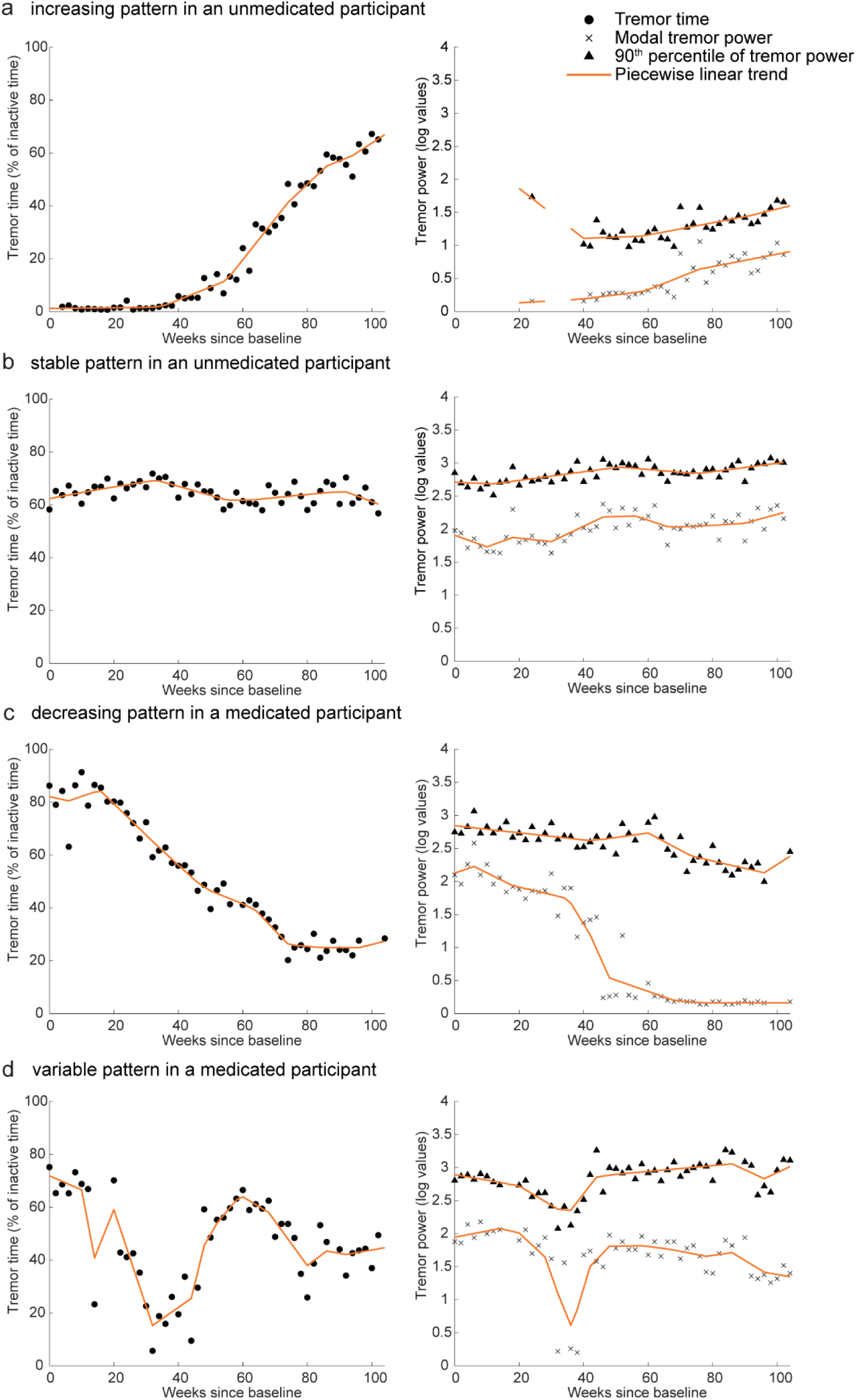
Examples of individual progression patterns in sensor-derived tremor measures. Tremor time is shown on the left, and the modal and 90^th^ percentile of tremor power in the graphs on the right. The two participants shown in **a** and **b** were not using symptomatic medication during the full study period, whereas the participants in **c** and **d** did. In **a** no tremor power measures were derived for the first weeks, since tremor time was below the false positive threshold of 3.5% in those weeks.

To compare the sensitivity to tremor progression between the sensor-derived tremor measures and clinical tremor scores, we derived SRMs at one- and two-year follow-ups. In the unmedicated tremor group (i.e., participants with tremor time above the false positive threshold), all sensor-derived tremor measures increased over two years (Figure 4). We observed a large responsiveness of tremor time, already after one year. The modal and 90^th^ percentile of tremor power demonstrated small-to-moderate responsiveness. In contrast to the sensitivity of sensor-derived measures, we observed no or small responsiveness of MDS-UPDRS tremor scores. The SRM for tremor time was significantly larger than those of all three MDS-UPDRS tremor items at one-year follow-up, and significantly larger than the rest tremor constancy (MDS-UPDRS 3.18) and patient-reported tremor score (MDS-UPDRS 2.10) after two years (differences and 95%-CI, computed using 10,000 bootstrap samples, are given in Supplementary Table 2). The 90^th^ percentile of tremor power also had a larger SRM at one-year follow-up compared to all three MDS-UPDRS tremor items.

**Figure 4:**
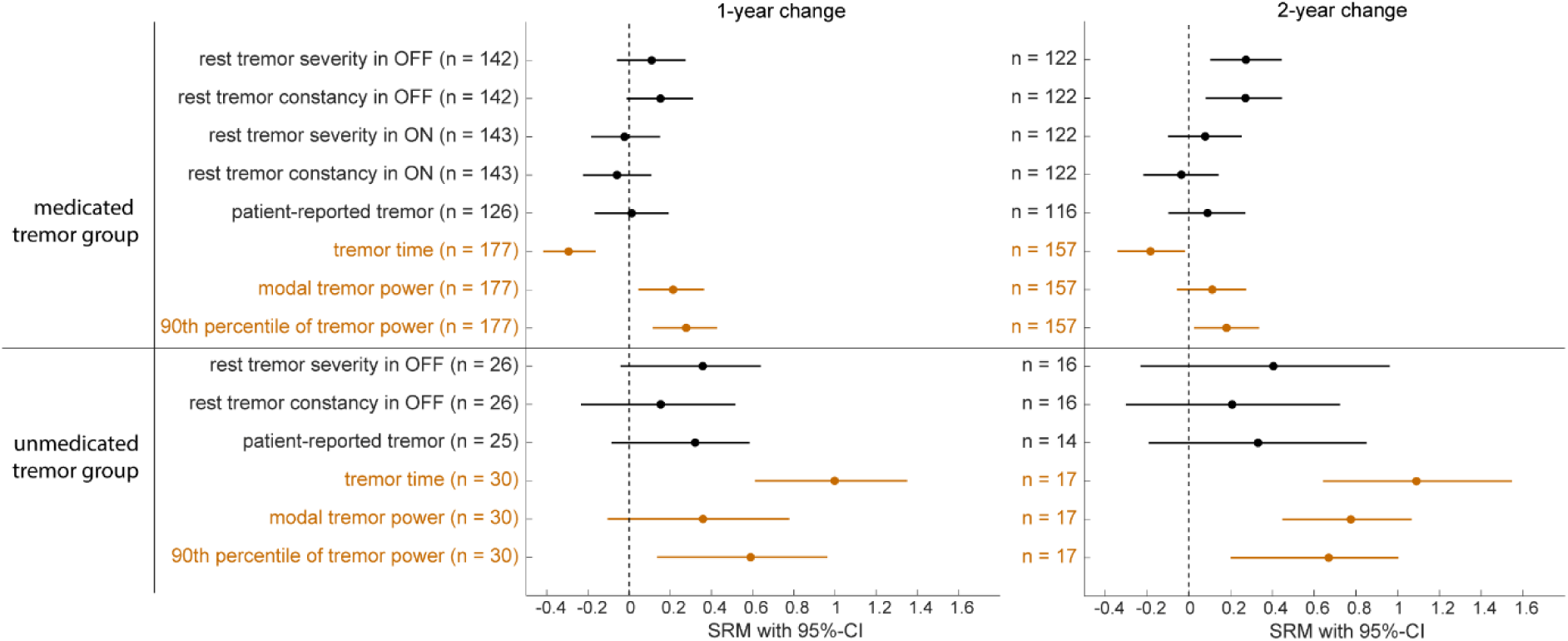
Standardized response mean (SRM) with 95% confidence interval (CI) of MDS-UPDRS tremor scores (black) and sensor-derived tremor measures (orange) assessed at one and two years after baseline. All participants with tremor time above the false positive threshold at baseline and at one- or two-year follow-up were included.

In the medicated tremor group, we observed a small improvement in tremor time (Figure 4). In contrast, the modal and 90^th^ percentile of tremor power slightly worsened. Regarding the sensitivity of clinical tremor scores, MDS-UPDRS ON state and patient-reported tremor scores did not show significant changes at one and two years of follow-up; sensor-derived tremor time was significantly more sensitive to improvement after one year compared to the clinical scores assessed in ON motor state and the patient-reported tremor score (Supplementary Table 2). The MDS-UPDRS OFF state assessments demonstrated slight worsening, comparable to the sensor-derived tremor power measures. The findings for tremor time and clinical tremor ratings were similar in the full unmedicated and medicated groups (including participants with and without tremor, see Supplementary Figure 3).

### Sensitivity analyses

To assess the impact of the follow-up duration on SRMs for the sensor-derived tremor measures in the two-year unmedicated (tremor) group, we visualized these over time in Supplementary Figure 4. A significant worsening of tremor time was already observed after 30 weeks, but for the tremor power measures this was only seen after 60-70 weeks.

Supplementary Figure 5 shows the SRMs based on the weekly tremor measures themselves (without estimating the piecewise linear trend) in the two-year unmedicated group. As expected, this assessment resulted in a more variable SRM over time. Although an increase in all three sensor-derived tremor measures is still visible, this analysis shows that smoothing is important to isolate long-term changes.

Reweighting the unmedicated group using IPCW had a small effect on the obtained SRMs, without changing the overall conclusions (see Supplementary Figure 6 for the unweighted results). Results of the Cox proportional hazards model fitted on participants unmedicated at baseline are shown in Supplementary Table 3.

Lastly, differences in tremor progression between the medicated and unmedicated groups could not be explained by differences in disease duration, age and watch side (see Supplementary Figure 7 for the SRMs obtained for the matched medicated group and Supplementary Table 4 for the baseline characteristics of this group).

### Correlation with patient-reported changes

We observed small but significant correlations with patient-reported tremor severity (MDS-UPDRS part II) for all sensor-derived tremor measures (tremor time: ρ = 0.17 (p<0.001; n=375); modal tremor power: ρ = 0.17 (p<0.05; n=144); 90^th^ percentile of tremor power: ρ = 0.19 (p<0.05; n=144). Regarding the MDS-UPDRS part III rest tremor scores, two-year changes in the rest tremor constancy score (MDS-UPDRS 3.18) and patient-reported tremor score were also weakly correlated, with Spearman’s ρ of 0.18 (p<0.001; n=387) for the OFF score and 0.19 (p<0.001; n=336) for the ON score. However, changes in the patient-reported tremor score did not significantly correlate with changes in the MDS-UPDRS rest tremor severity scores, both in ON (Spearman’s ρ of 0.04; p=0.11; n=336) and OFF (Spearman’s ρ of 0.08; p=0.42; n=387).

### Effect of changes in dopaminergic medication dose

We assessed the association between two-year changes in dopaminergic medication dose and daily-life tremor measures in the medicated group, adjusted for disease duration and watch side. A larger increase in LEDD over the two-year period was weakly associated with a greater reduction in tremor time (standardized β=-0.10, 95%-CI -0.21 – 0.01). In addition, a higher disease duration at baseline was associated with a greater reduction in tremor time (standardized β=-0.15, 95%-CI -0.26 – -0.03). Changes in the tremor power measures were not associated with changes in the studied clinical factors (see Supplementary Figure 8).

The responsiveness to dopaminergic treatment initiation of tremor time (n=39) and of the modal and 90^th^ percentile of tremor power (n=23) are shown in Figure 5. Tremor time decreased after the initiation of treatment, whereas the tremor power measures did not show a significant decrease.

**Figure 5:**
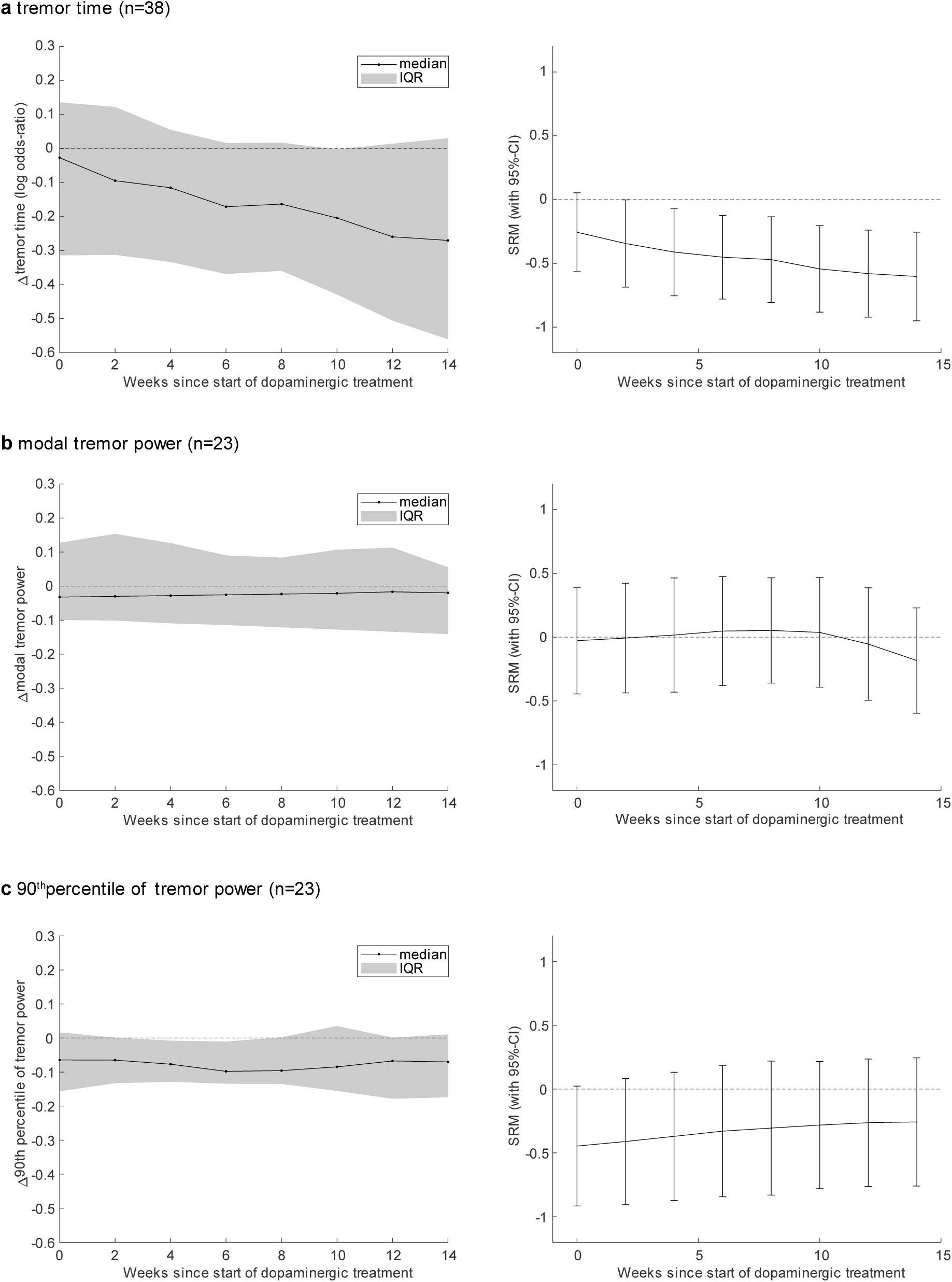
Responsiveness to the initiation of dopaminergic treatment of a) tremor time, b) modal tremor power, and c) the 90^th^ percentile of tremor power. The absolute changes in tremor measures are shown on the left. From these changes the standardized response means (SRMs) were computed, shown on the right.

## Discussion

Using wrist sensor data obtained from two years of passive monitoring, we demonstrated that prior to symptomatic drug treatment initiation, daily-life sensor-derived measures are more sensitive to tremor progression than both clinician- and patient-reported outcomes. In particular, sensor-derived tremor time demonstrated a large responsiveness to tremor progression at both one and two years of follow-up. By contrast, in medicated participants, tremor time significantly decreased over time, which was associated with both a longer disease duration and increasing dopaminergic medication dose (i.e., LEDD). Together, these findings show that sensor-derived tremor time is a sensitive digital biomarker for quantifying both tremor progression and treatment response, with promising applications in clinical trials and individual patient care.

Sensitive outcome measures are critical for quantifying disease progression in clinical trials, not only for PD but also for other chronic diseases. Many new disease-modifying therapies are being developed, but evaluating their efficacy currently requires large sample sizes (typically over 200 patients) and long follow-up^32^. In the unmedicated group, which is the target population for many disease-modifying trials, tremor time already increased over one year of follow-up, with an SRM approximately twice as large compared to those of MDS-UPDRS rest tremor scores. This implies that, by using passive digital monitoring of tremor instead of clinically based tremor assessments, a four times smaller sample size is required in a typical randomized controlled trial design^33^. The one-year SRM for tremor time that we observed in the full unmedicated group (0.65) is equal to that observed in the WATCH-PD study^7^. In unmedicated participants with tremor, we observed an even larger SRM (1.09 after one year), which was similar after one and two years of follow-up.

In the medicated group, we observed a small but significant improvement in tremor time, while the 90^th^ percentile of tremor power slightly worsened over one and two years. Furthermore, a decrease in tremor time was associated with an increase in LEDD, whereas this association was absent for the 90^th^ percentile of tremor power. Based on these findings, it is likely that the 90^th^ percentile of tremor power reflects the tremor severity during OFF periods in daily life, and is therefore more sensitive to capture disease progression in medicated participants. This aligns with findings from the clinical tremor assessments, showing that tremor severity and constancy in OFF also worsened over time in the medicated group, in agreement with previous studies^17–19^. By contrast, weekly tremor time is likely more influenced by ON than OFF periods, since early-stage PD participants on symptomatic medication typically spend most of their time in the ON state^34^. Therefore, the observed improvements in tremor time can be partly explained by up-titration of symptomatic medication in early-stage PD. Noteworthy, improvements in tremor time were not only related to increases in LEDD in the medicated group, but also to increased disease duration. This aligns with previous findings that tremor increases in the early stages of PD but can disappear later in the disease (inverted u-shape hypothesis of tremor progression^20,35,36^). This is important to consider in clinical trials, since the largest treatment effects are expected to be found in participants with short disease duration.

An important requirement for using sensor-derived measures in clinical trials and clinical care is the ability to capture meaningful changes^9,10^. Digital measures of tremor have been identified as relevant by persons with early PD^37^, but showing the ability to capture meaningful changes remains difficult. Although we previously observed moderate correlations between sensor-derived tremor measures and the patient-reported tremor score cross-sectionally^23^, here we observed small but significant correlations longitudinally. This may be partly explained by the limited sensitivity to change of episodic patient-reported tremor scores^4^. A more suitable anchor to determine meaningful change might be the Patient Global Impression of Change scale (PGI-C)^38^. By assessing both the PGI-C score and the change in sensor-derived tremor measures after an intervention where change is expected, the minimal clinically important differences of the tremor measures could be determined.

## Strengths and limitations

A key strength of this study is the use of a unique large dataset of continuous wrist sensor data, collected over a two-year period with minimal drop-out and high wearing compliance^24^. Second, in contrast to proprietary algorithms used in other studies, we applied a validated, open-source and device-agnostic algorithm to derive daily-life tremor measures, ensuring that our results are directly relevant for other studies collecting wrist gyroscope data. Third, we conducted in-depth analyses regarding the performance of sensor-derived tremor measures in both unmedicated and medicated groups of early PD. This could serve as an example for evaluating digital biomarkers in future research, for PD as well as for other chronic diseases. Finally, to obtain unbiased estimates of tremor progression in the unmedicated group, we corrected for informative censoring due to earlier treatment initiation in participants with more severe tremor. Our approach could be used in disease-modifying clinical trials as well, which typically also censor patients after initiation of symptomatic treatment, but often do not account for informative censoring^39^.

A limitation of this study is that LEDD information was only yearly available, so that the effect of changes in LEDD on daily-life tremor progression could not be assessed on a more granular week-to-week basis. Furthermore, the effect of dopaminergic treatment on tremor is highly variable, which could explain the weak negative relation between change in tremor time and change in LEDD^40^. It is also possible that a more aggressive worsening of tremor over time necessitated a larger increase in LEDD; the net result is a weak correlation between tremor worsening and LEDD because these two effects oppose each other.

Another important consideration is that we focussed only on rest (and re-emergent) tremor. Although this is the most characteristic and prevalent tremor phenotype in PD, kinetic and pure postural tremor may have different progression patterns^20^. Lastly, although the use of a two-year unmedicated group of PD participants is unique, the sample size of this subgroup was small because many participants initiated symptomatic therapy. The results of this study should therefore be replicated and validated in independent cohorts. Nevertheless, the findings on tremor progression in unmedicated PD are directly relevant to disease - modifying clinical trials, as these focus increasingly on de novo PD or even prodromal disease stages^41^. Tremor is the most common presenting symptom at disease onset, and has been observed in prodromal cases^35,42–44^. Future research could explore whether real-life detection of tremor in prodromal PD is possible as well^45^.

In conclusion, this study shows the potential of daily-life sensor-derived tremor measures for evaluating novel treatments in clinical trials. Our findings also contribute new insights into daily-life tremor progression in PD. The sensor-derived tremor measures could be combined with other biomarkers in future studies, such as neuroimaging markers, to improve understanding of individual variation in tremor expression. Lastly, although tremor is a highly relevant symptom for persons with PD, other motor and non-motor symptoms affect their quality of life as well^12^. Future research should aim to combine real-life monitoring of tremor with monitoring of other relevant symptoms in PD.

## Author contributions

NAT, IGB, RCH and LJWE conceptualized the study. DCS, EP, MAL and LJWE contributed to the methodology. NAT performed the data analysis, created the figures and wrote the original draft. IGB, BRB, RCH and LJWE supervised the study. IGB, DCS, EP, HC, SS, MAL, YPR, BRB, RCH and LJWE reviewed and edited the manuscript. HC, SS, MAL, BRB and LJWE were involved in funding acquisition. NAT directly accessed and verified the underlying data. All authors had full access to all the data in the study and had final responsibility for the decision to submit for publication.

## Data availability

Data from the Personalized Parkinson Project used in the present study were retrieved from the PEP database (https://pep.cs.ru.nl/index.html). The PPP data is available upon request via. More details on the procedure can be found on the website www.personalizedparkinsonproject.com/home. The code used to conduct this study is publicly available at https://github.com/AI-for-Parkinson-Lab/longitudinal_tremor.

## Declaration of interests

S.S. is currently employed by, and currently holds shares in, Verily Life Sciences, but declares no nonfinancial competing interests. B.R.B. serves as the co-Editor in Chief for the Journal of Parkinson’s disease, serves on the editorial board of Practical Neurology and Digital Biomarkers, has received fees from serving on the scientific advisory board for the Critical Path Institute, Gyenno Science, MedRhythms, UCB, Kyowa Kirin and Zambon (paid to the Institute), has received fees for speaking at conferences from AbbVie, Bial, Biogen, GE Healthcare, Oruen, Roche, UCB and Zambon (paid to the Institute), and has received research support from Biogen, Cure Parkinson’s, Davis Phinney Foundation, Edmond J. Safra Foundation, Fred Foundation, Gatsby Foundation, Hersenstichting Nederland, Horizon 2020, IRLAB Therapeutics, Maag Lever Darm Stichting, Michael J Fox Foundation, Ministry of Agriculture, Ministry of Economic Affairs & Climate Policy, Ministry of Health, Welfare and Sport, Netherlands Organization for Scientific Research (ZonMw), Not Impossible, Parkinson Vereniging, Parkinson’s Foundation, Parkinson’s UK, Stichting Alkemade-Keuls, Stichting Parkinson NL, Stichting Woelse Waard, Health Holland / Topsector Life Sciences and Health, UCB, Verily Life Sciences, Roche and Zambon. B.R.B. does not hold any stocks or stock options with any companies that are connected to Parkinson’s disease or to any of his clinical or research activities. All other authors declare no competing interests.

## Acknowledgments

We would like to thank all Personalized Parkinson Project participants, assessors and project coordinators, who made it possible to collect the unique dataset of continuous wearable sensor data used in this study. This work was financially supported by the Michael J Fox Foundation (grant #020425), the Dutch Research Council (grant #ASDI.2020.060 & grant #2023.010), and the Dutch Research Council Long-Term Program (project #KICH3.LTP.20.006, financed by the Dutch Research Council, Verily, and the Dutch Ministry of Economic Affairs and Climate Policy).The Personalized Parkinson Project is co-funded by Verily Life Sciences LLC, Radboud University Medical Center and by the PPP Allowance made available by Health∼Holland, Top Sector Life Sciences & Health, to stimulate public-private partnerships. H.C. received research grants from the UK Medical Research Council (grant #MR/R020418/1 & grant #MR/X023141/1). I.G.B. was financially supported via the Radboud Healthy Data program. R.C.H. received research grants from the MJ Fox Foundation, the Netherlands Brain Foundation, ParkinsonNL, and the Netherlands Organization for Health Research and Development.

## Notes

### Author Declarations

The study received approval from the Commissie Mensgebonden Onderzoek Region Arnhem-Nijmegen, and all participants provided informed consent.

