## Supplementary material for "Progression of daily-life tremor measures in early Parkinson disease: a longitudinal continuous monitoring study"

*Supplementary Table 1: The optimal λ used for estimating the piecewise linear trends and subsequently derived proportion of variance explained by the fitted trends of sensor-derived tremor measures. The variances were derived for medicated and unmedicated participants separately. IQR = interquartile range.*

| | **λ** | **Proportion of variance explained by the fitted trends in medicated participants – median (IQR)** | **Proportion of variance explained by the fitted trends in unmedicated participants – median (IQR)** |
|---|---|---|---|
| **Tremor time** | 0.09 | 0.34 (0.15 – 0.61) | 0.36 (0.12 – 0.69) |
| **Modal tremor power** | 0.35 | 0.38 (0.21 – 0.62) | 0.47 (0.25 – 0.60) |
| **$90^{th}$ percentile of tremor power** | 0.56 | 0.43 (0.22 – 0.63) | 0.43 (0.27 – 0.73) |

*Supplementary Table 1: Differences (and 95% confidence intervals, assessed using bootstrapping) between standardized response means (SRMs) of sensor-derived tremor measures and MDS-UPDRS tremor scores, for the different subgroups and follow-up periods used in the analyses. In the medicated groups, we compared the SRMs of tremor time to clinical rest tremor scores assessed in ON, whereas we compared the SRMs of the modal and $90^{th}$ percentile of tremor power to those of OFF clinical scores. Significant differences are highlighted in bold.*

| | | | **Rest tremor severity** | **Rest tremor constancy** | **Patient-reported tremor** |
|---|---|---|---|---|---|
| Unmedicated tremor group | One-year change | Tremor time | **0.58 (0.09 – 1.12)**<br>n=26 | **0.79 (0.27 – 1.34)**<br>n=26 | **0.63 (0.14 – 1.19)**<br>n=25 |
| | | Modal tremor power | 0.13 (-0.44 – 0.74)<br>n=26 | 0.34 (-0.27 – 0.98)<br>n=26 | 0.19 (-0.39 – 0.81)<br>n=25 |
| | | $90^{th}$ percentile of tremor power | **0.53 (0.07 – 1.04)**<br>n=26 | **0.74 (0.24 – 1.28)**<br>n=26 | **0.67 (0.21 – 1.18)**<br>n=25 |
| | Two-year change | Tremor time | 0.69 (-0.08 – 1.48)<br>n=16 | **0.94 (0.25 – 1.72)**<br>n=16 | **0.78 (0.01 – 1.63)**<br>n=14 |
| | | Modal tremor power | 0.36 (-0.34 – 1.04)<br>n=16 | 0.59 (-0.00 – 1.25)<br>n=16 | 0.46 (-0.22 – 1.17)<br>n=14 |
| | | $90^{th}$ percentile of tremor power | 0.34 (-0.42 – 1.14)<br>n=16 | 0.58 (-0.07 – 1.38)<br>n=16 | 0.68 (-0.19 – 1.67)<br>n=14 |
| Unmedicated group | One-year change | Tremor time | 0.25 (-0.09 – 0.61)<br>n=39 | **0.52 (0.14 – 0.91)**<br>n=39 | **0.56 (0.12 – 0.97)**<br>n=37 |
| | Two-year change | Tremor time | 0.20 (-0.39 – 0.81)<br>n=23 | 0.43 (-0.06 – 0.99)<br>n=23 | **0.67 (0.05 – 1.34)**<br>n=19 |
| Medicated tremor group | One-year change | Tremor time | **-0.31 (-0.54 - -0.08)**<br>n=143 | **-0.27 (-0.50 - -0.04)**<br>n=143 | **-0.37 (-0.61 - -0.13)**<br>n=126 |
| | | Modal tremor power | -0.12 (-0.13 - 0.37)<br>n=142 | 0.07 (-0.17 - 0.33)<br>n=142 | 0.22 (-0.04 - 0.49)<br>n=126 |
| | | $90^{th}$ percentile of tremor power | 0.19 (-0.04 - 0.43)<br>n=142 | 0.15 (-0.09 - 0.38)<br>n=142 | **0.27 (0.02 - 0.52)**<br>n=126 |
| | Two-year change | Tremor time | -0.22 (-0.47 – 0.03)<br>n=122 | -0.11 (-0.37 – 0.14)<br>n=122 | -0.24 (-0.50 – 0.02)<br>n=116 |
| | | Modal tremor power | -0.11 (-0.36 – 0.13)<br>n=122 | -0.11 (-0.37 – 0.14)<br>n=122 | 0.05 (-0.21 – 0.31)<br>n=116 |
| | | $90^{th}$ percentile of tremor power | -0.09 (-0.34 – 0.16)<br>n=122 | -0.09 (-0.34 – 0.17)<br>n=122 | 0.06 (-0.21 – 0.31)<br>n=116 |
| Medicated group | One-year change | Tremor time | **-0.34 (-0.48 - -0.20)**<br>n=378 | **-0.28 (-0.42 - -0.14)**<br>n=378 | **-0.28 (-0.44 - -0.13)**<br>n=338 |
| | Two-year change | Tremor time | **-0.25 (-0.40 - -0.10)**<br>n=357 | -0.11 (-0.25 - 0.04)<br>n=357 | -0.11 (-0.27 - 0.05)<br>n=329 |

*Supplementary Table 2: Results of the Cox proportional hazards model fitted on participants unmedicated at baseline (n=78), to estimate the effect of tremor on the time to treatment initiation. Different lags (4, 6 and 8 weeks) between the measurement of tremor time and the initiation of treatment were evaluated in separate models. The model with a lag of 4 weeks was selected based on the largest concordance statistic, although the difference between the three models was small.*

| Lag between tremor time measurement week and initiation of treatment (number of weeks) | Beta coefficient in Cox proportional hazards model (95%-CI) | Concordance statistic |
|---|---|---|
| 4 | 0.89 (-0.11 – 1.50) | 0.535 |
| 6 | 0.83 (-0.17 – 1.51) | 0.531 |
| 8 | 0.80 (-0.20 – 1.51) | 0.528 |

*Supplementary Table 4: Demographic and clinical characteristics, as well as sensor-derived tremor measures at baseline of the unmedicated and matched medicated groups. The modal and 90$^{th}$ percentile of tremor power were assessed in all participants with tremor time above the false positive threshold at baseline (n=47 for the unmedicated group and n=38 for the matched medicated group). IQR: inter-quartile range. MDS-UPDRS: Movement Disorder Society-Sponsored Revision of the Unified Parkinson's Disease Rating Scale. Part 1: non-motor experiences of daily living. Part 2: motor experiences of daily living. Part 3: motor examination. Part 4: motor complications. * Tremor power measures were only derived for participants with tremor time above the false positive threshold (n=47 for the unmedicated group and n=38 for the matched medicated group).*

| | | Unmedicated group (n=78) | Matched medicated group (n=78) |
|---|---|---|---|
| **Demographic and clinical characteristics at baseline** | Age (years), median (IQR) | 65 (56 – 70) | 62 (55 – 69) |
| | Gender (men), n (%) | 48 (62) | 45 (58) |
| | Disease duration (months), median (IQR) | 5 (2 – 13) | 10 (7 – 14) |
| | Watch worn on more-affected side, n (%) | 60 (78) | 58 (74) |
| | MDS-UPDRS part 1, median (IQR) | 5 (3 – 7) | 9 (6 – 13) |
| | MDS-UPDRS part 2, median (IQR) | 4 (3 – 8) | 4 (2 – 9) |
| | MDS-UPDRS part 3 OFF, median (IQR) | 34 (28 – 40) | 28 (19 – 37) |
| | MDS-UPDRS part 4, median (IQR) | 0 (0 – 1) | 0 (0 – 4) |
| | MDS-UPDRS part 3 OFF tremor subscore, median (IQR) | 5 (2 – 9) | 4 (2 – 6) |
| | Rest tremor severity device-sided arm, median (IQR) | 1 (0 – 2) | 0 (0 -1) |
| **Sensor-derived tremor measures at baseline** | Tremor time (%), median (IQR) | 5.8 (1.4 – 43.1) | 2.8 (1.1 – 10.3) |
| | Modal tremor power (log values), median (IQR)* | 0.7 (0.2 – 1.2) | 0.3 (0.2 – 0.5) |
| | 90$^{th}$ percentile of tremor power (log values), median (IQR)* | 1.9 (1.5 – 2.3) | 1.5 (1.2 – 1.9) |

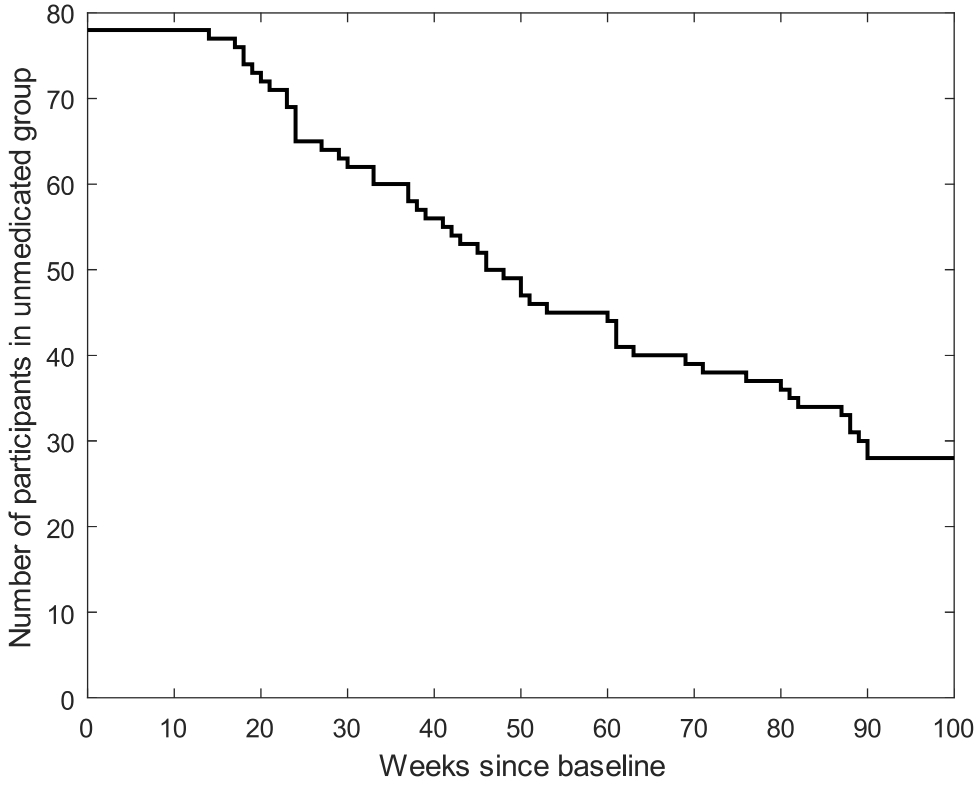


*Supplementary Figure 1: Number of participants included in the unmedicated group, which decreased over time due to the initiation of symptomatic drug treatment in some participants.*

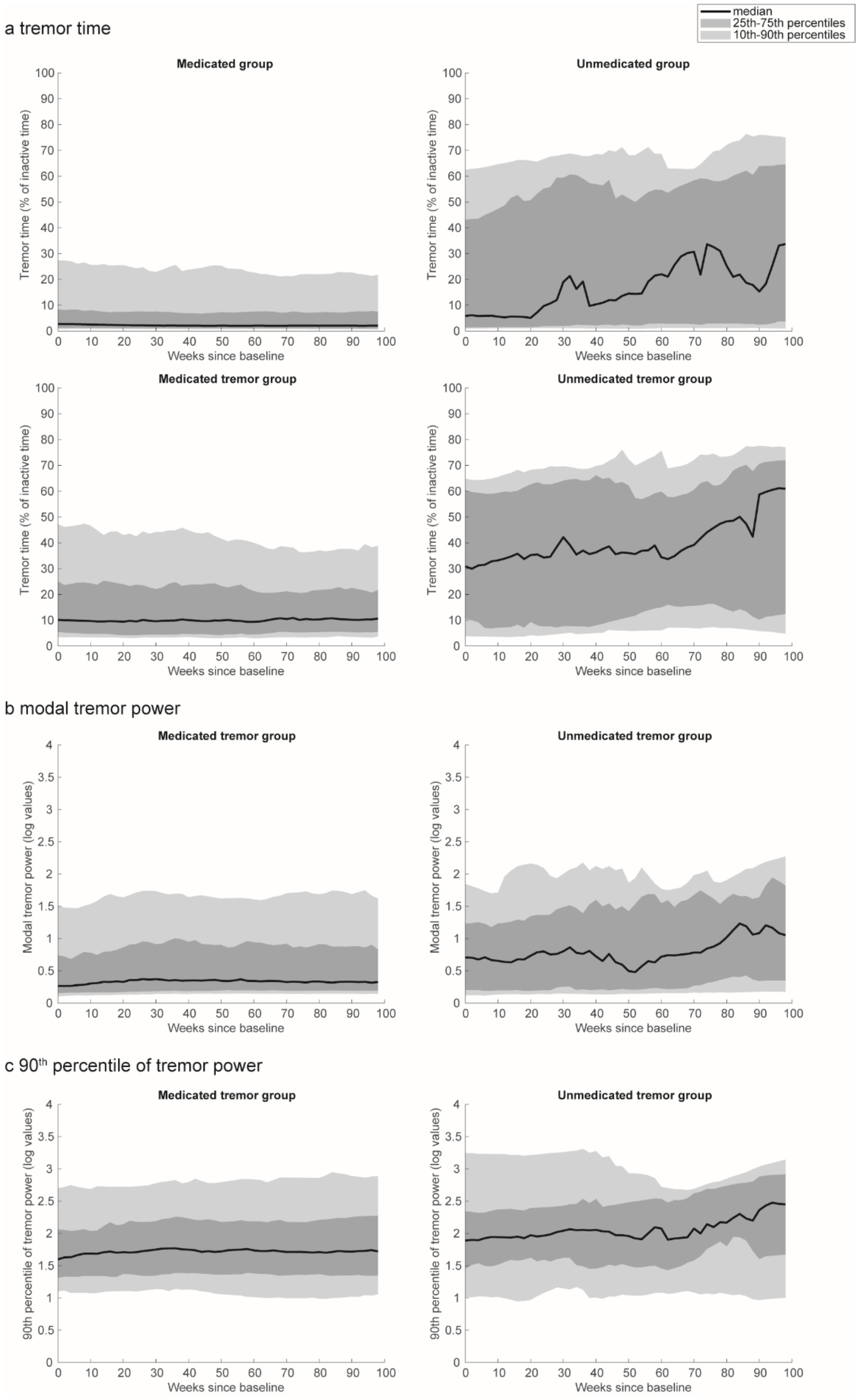


*Supplementary Figure 2: Distribution of sensor-derived tremor measures over time in the medicated group (on the left) and unmedicated group (on the right). Tremor time (a) was derived for all participants and weeks in which data was available (upper plots), and subsequently in all participants and weeks in which tremor time was above the false positive threshold (lower plots). The modal (b) and 90$^{th}$ percentile of tremor power (c) were also assessed for the participants and weeks in which tremor time was above the false positive threshold.*

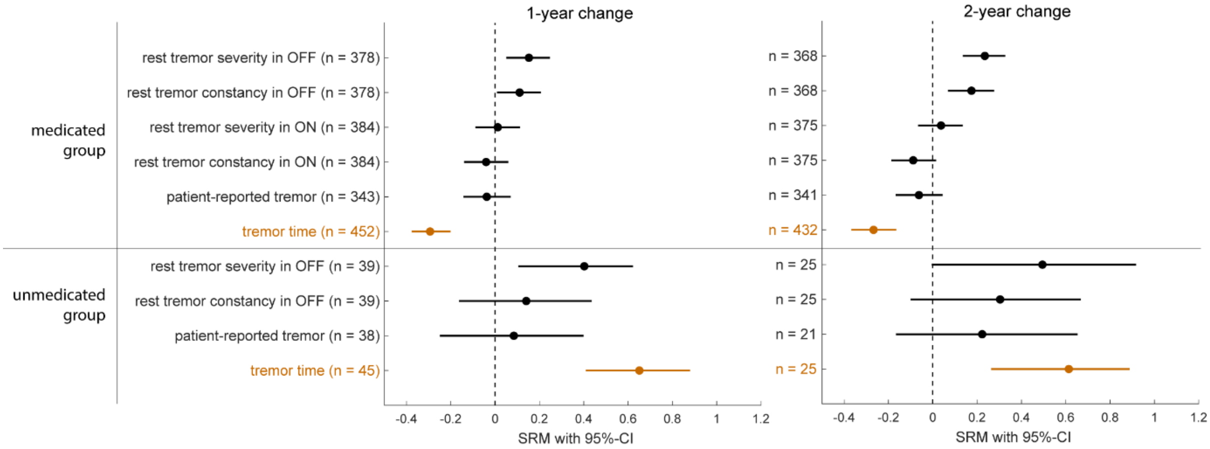


*Supplementary Figure 3: Standardized response mean (SRM) with 95% confidence interval (CI) of MDS-UPDRS tremor scores (black) and sensor-derived tremor measures (orange) assessed at one and two years after baseline. All participants with data available at baseline and at one- or two-year follow-up were included.*

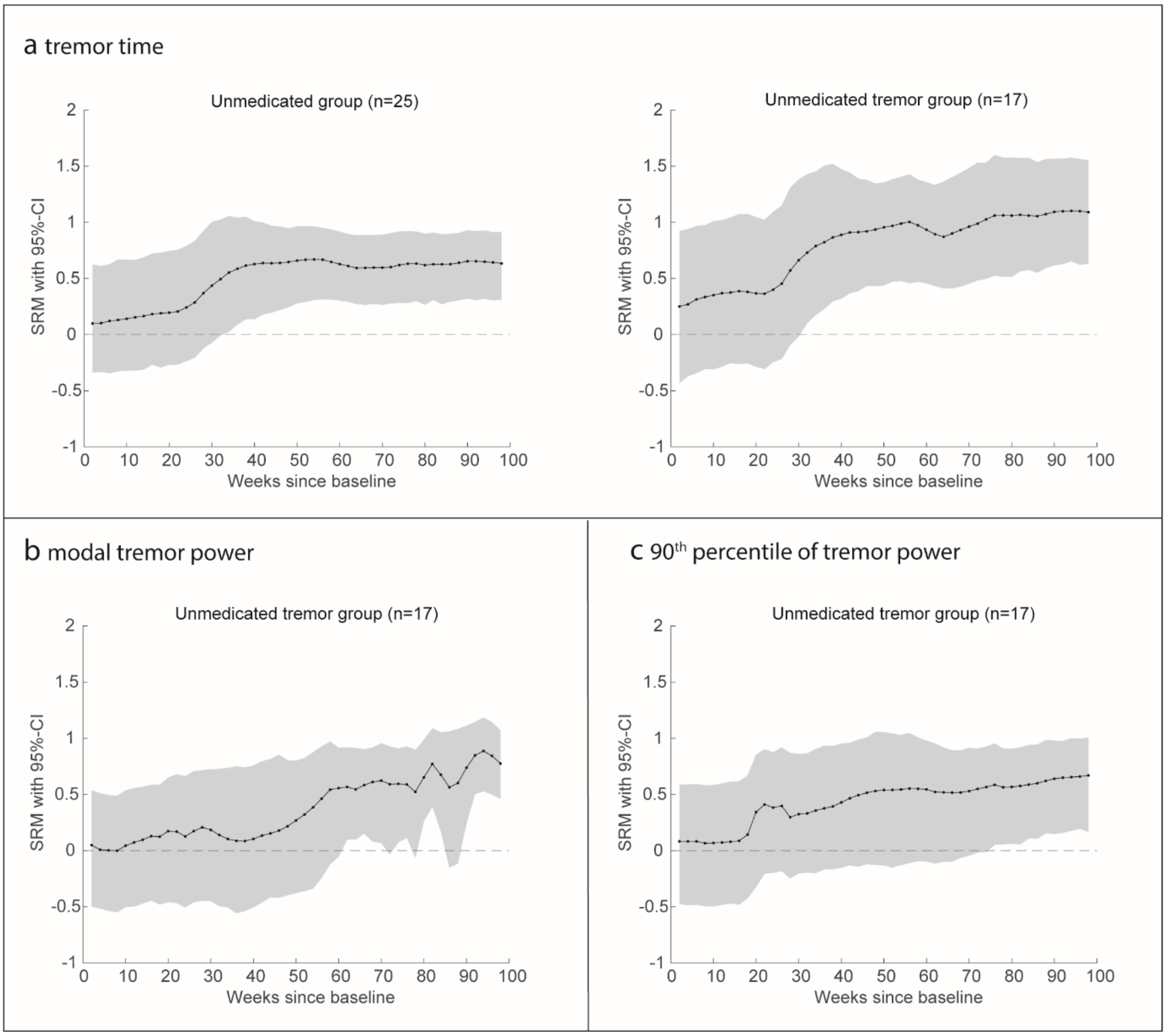


*Supplementary Figure 4: Standardized response mean (SRM) with 95% confidence interval (CI) of sensor-derived tremor measures over time in the two-year unmedicated group. Tremor time (a) was assessed in all participants with data available at baseline and at two-year follow-up (left), and subsequently in all participants with tremor time above the false positive threshold at baseline and two-year follow-up (right). The modal (b) and 90$^{th}$ percentile of tremor power (c) were also assessed in all participants with tremor time above the false positive threshold at baseline and at two-year follow-up.*

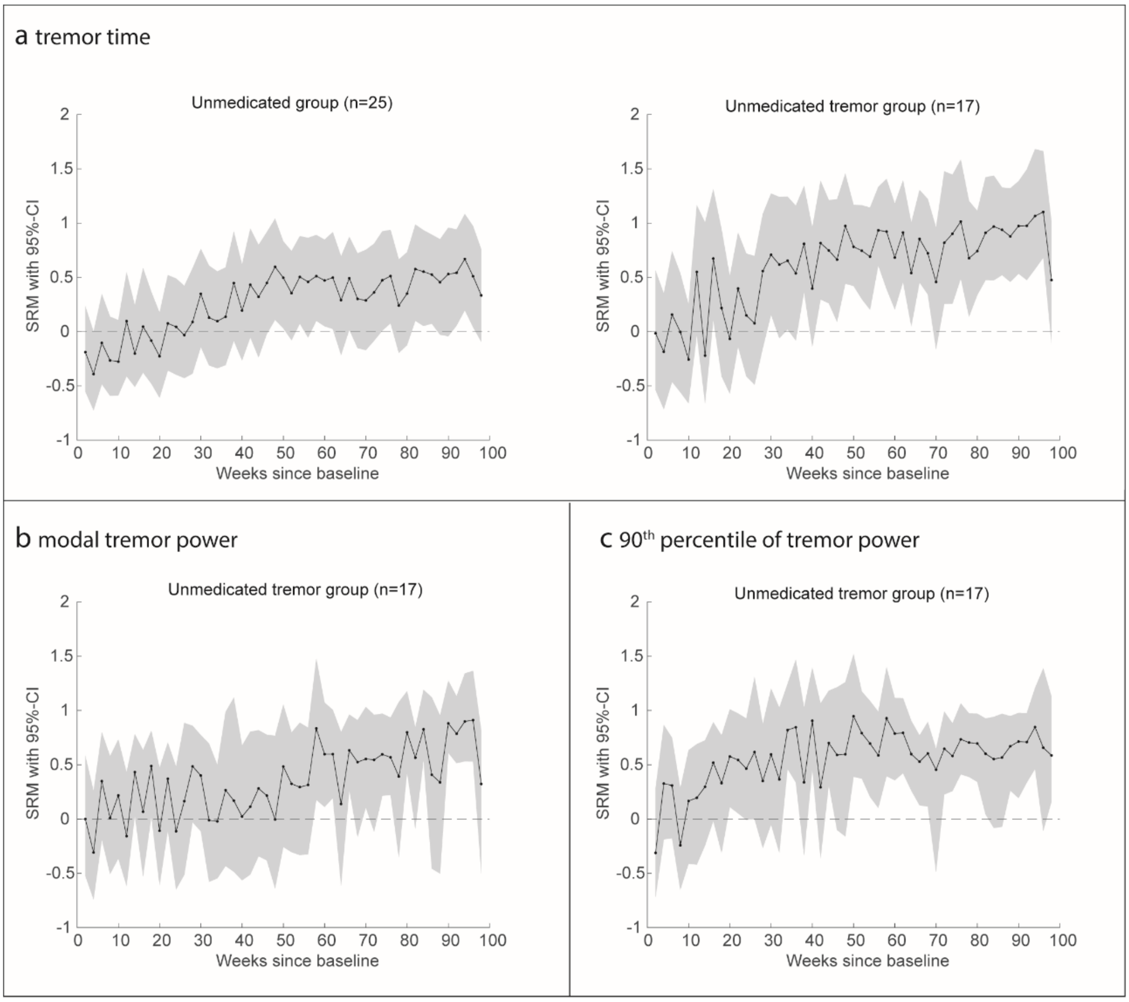


*Supplementary Figure 5: Standardized response mean (SRM) with 95% confidence intervals (CI) of sensor-derived tremor measures over time, based on weekly tremor measures assessed in the two-year unmedicated group (without piecewise linear trend estimation). Tremor time (a) was assessed in all participants with data available at baseline and at two-year follow-up (on the left), and subsequently in all participants with tremor time above the false positive threshold at baseline and at two-year follow-up (on the right). The modal (b) and 90$^{th}$ percentile of tremor power (c) were also assessed in all participants with tremor time above the false positive threshold at baseline and at two-year follow-up.*

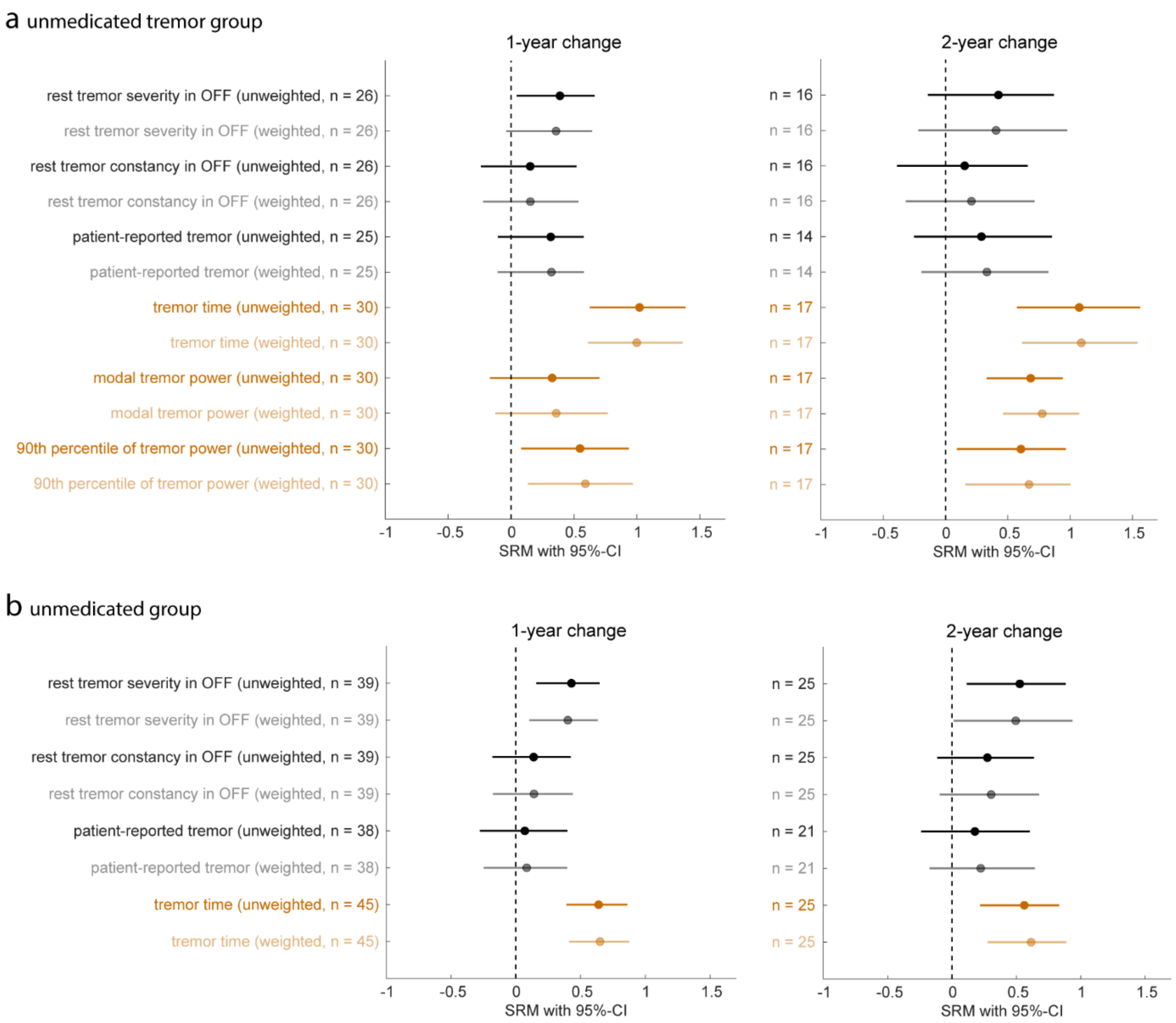


*Supplementary Figure 6: Standardized response mean (SRM) with 95% confidence interval (CI) of MDS-UPDRS tremor scores (black) and sensor-derived tremor measures (orange) assessed at one and two years after baseline in the weighted and unweighted unmedicated (tremor) groups.*

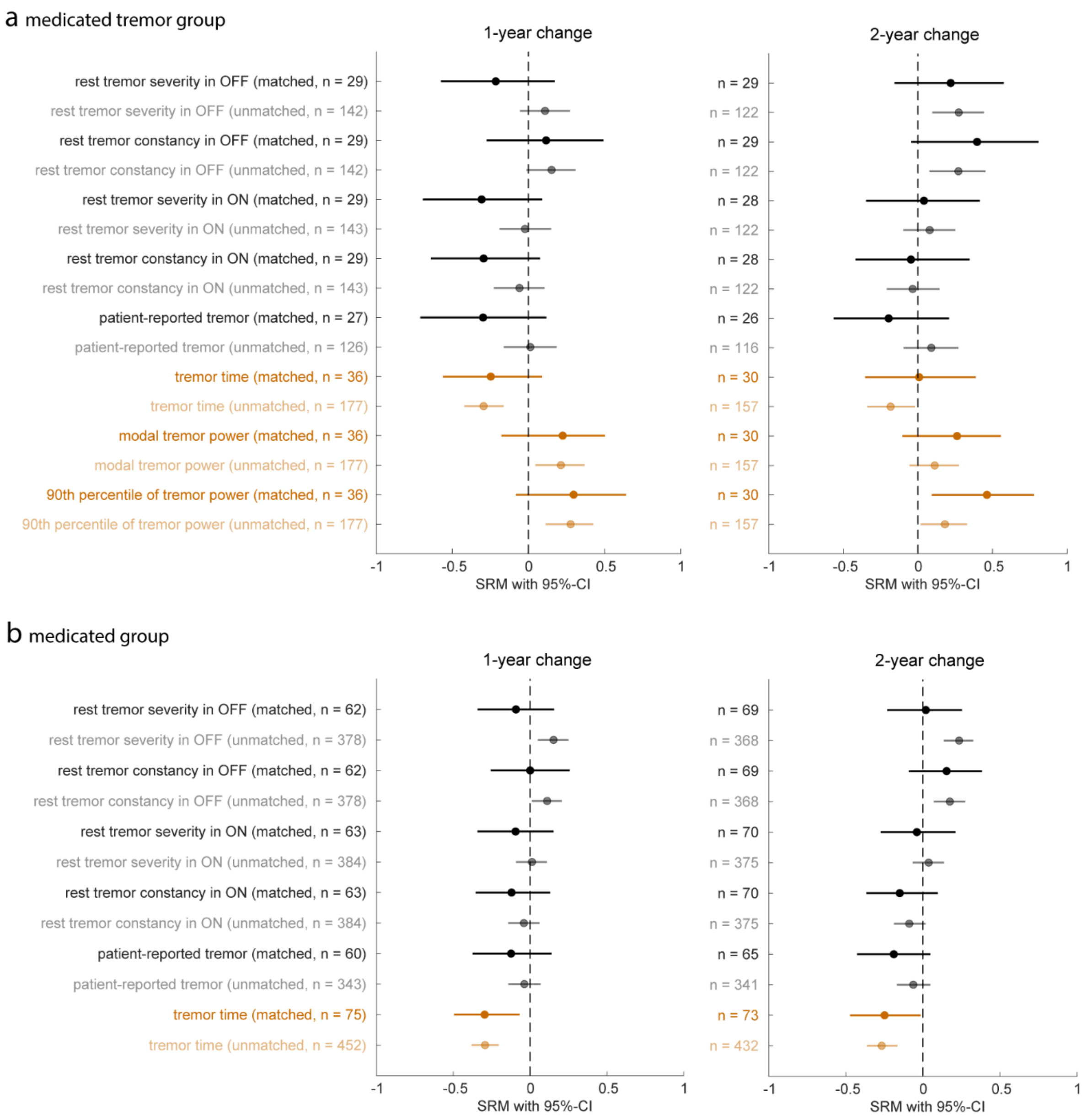


*Supplementary Figure 7: Standardized response mean (SRM) with 95% confidence interval (CI) of MDS-UPDRS tremor scores (black) and sensor-derived tremor measures (orange) assessed at one and two years after baseline, with and without matching the medicated group on the unmedicated group at baseline.*

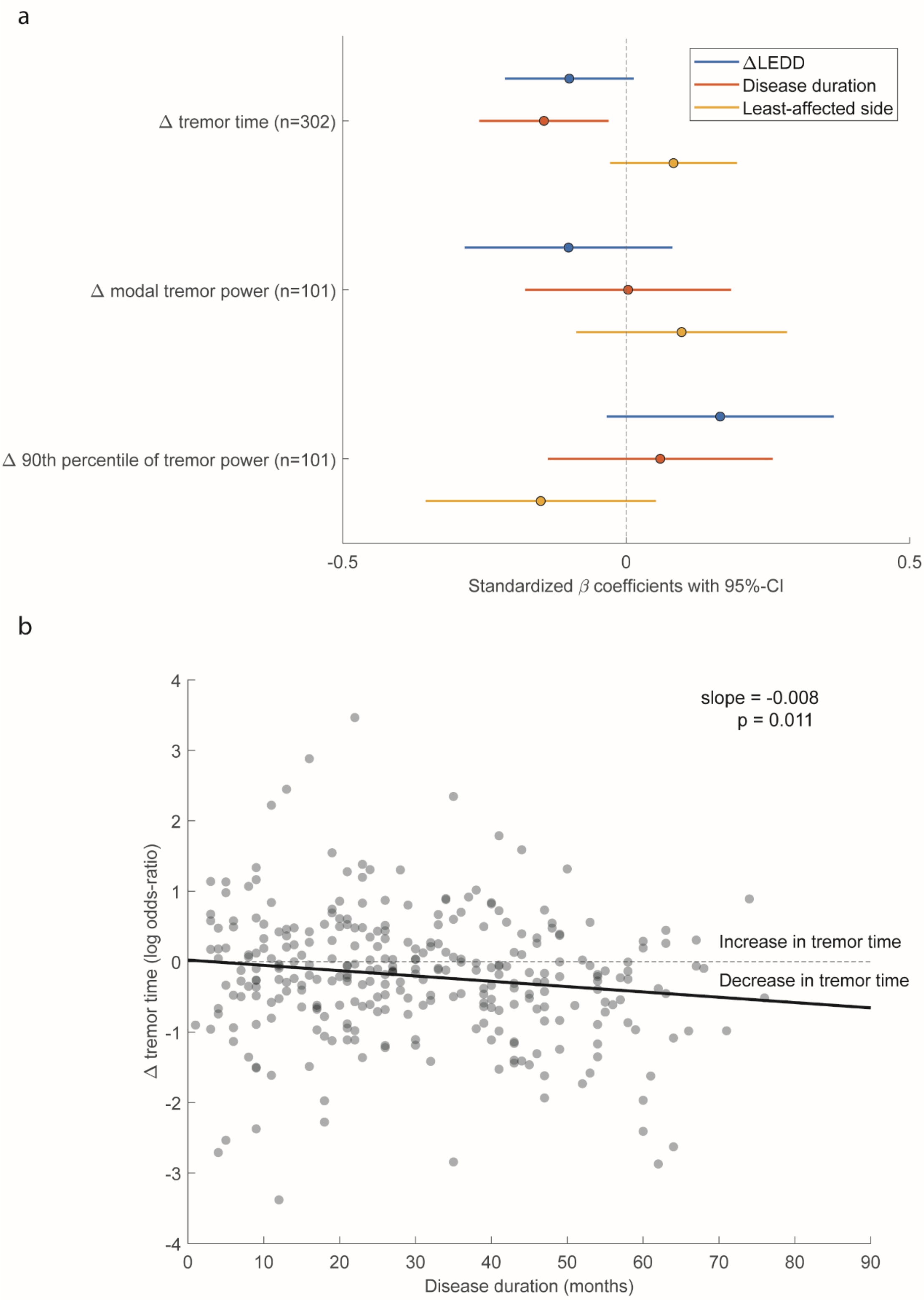


*Supplementary Figure 8: **a** shows the standardized β coefficients obtained by multivariable linear regression with two-year changes in sensor-derived tremor measures as individual outcomes and the following predictors: 1) changes in levodopa equivalent daily dose (LEDD), 2) disease duration at baseline and 3) watch side (more- or less-affected side). In **b** the individual two-year changes in tremor time against disease duration at baseline are shown, corrected for change in LEDD and taken as if the watch was worn on the more-affected side.*
